# Superior immunogenicity and effectiveness of the 3rd BNT162b2 vaccine dose

**DOI:** 10.1101/2021.12.19.21268037

**Authors:** Yaniv Lustig, Tal Gonen, Lilac Melzer, Mayan Gilboa, Victoria Indenbaum, Carmit Cohen, Sharon Amit, Hanaa Jaber, Ram Doolman, Keren Asraf, Carmit Rubin, Ronen Fluss, Ella Mendelson, Laurence Freedman, Gili Regev-Yochay, Yitshak Kreiss

**Affiliations:** Central Virology Laboratory, Public Health Services, Ministry of Health, Sheba Medical Center, Tel-Hashomer, Israel; Sackler Faculty of Medicine, Tel-Aviv University, Israel; The Infection Prevention & Control Unit, Sheba Medical Center, Tel Hashomer, Israel; Clinical Microbiology, Sheba Medical Center; Biostatistics and Biomathematics Unit, Gertner Institute of Epidemiology and Health Policy Research, Sheba Medical Center, Tel Hashomer, Israel; Automated Mega-Laboratory, Laboratory division, Sheba Medical Center; The Sheba Medical Center Management, Sheba Medical Center, Tel Hashomer, Israel

**Keywords:** BNT162 vaccine, COVID-19, immunogenicity, humoral response, IgG, neutralizing antibody, Third vaccine dose, safety, vaccine effectiveness

## Abstract

In a prospective cohort study involving 12,413 Health Care Workers (HCW), we assessed immunogenicity, vaccine-effectiveness (VE) and safety of the third BNT162b2 vaccine dose. One month after third dose, anti-RBD-IgG were induced 1.7-folds compared to one month after the second. A significant increase in avidity from 61.1% (95%CI:56.1-66.7) to 96.3% (95%CI:94.2-98.5) resulted in a 6.1-folds neutralizing antibodies induction. Linear mixed model demonstrated that the third dose elicited a greater response among HCW≥60 or those with ≥two comorbidities who had a lower response following the second dose. VE of the third dose relative to two doses was 85.6% (95% CI, 79.2-90.1%). No serious adverse effects were reported. These results suggest that the third dose is superior to the second dose in both quantity and quality of IgG-antibodies and safely boosts protection from SARS-CoV-2 infection by generating high avidity antibodies to levels that are not significantly different between healthy and vulnerable populations.

---

The vaccination campaign against coronavirus disease 2019 (COVID-19) is expanding worldwide and studies demonstrate that all FDA approved vaccines, and in particular mRNA-based BNT162b2 and mRNA-1273, show high vaccine efficacy and effectiveness in preventing symptomatic COVID-19 ^1-6^.

Disturbingly, COVID-19 breakthrough infections are substantially increasing, especially in countries which were of the first ones to initiate vaccine rollout. Accumulating evidence suggest that vaccine effectiveness is declining in all age groups, a few months after receipt of the second dose of vaccine^3,7,8^ and that humoral immunity follows a similar path^9^. These findings along with evidence that SARS-CoV-2 breakthrough infections are correlated with lower IgG and neutralizing antibody levels^10^, prompted Israeli authorities to approve the administration of a third vaccine dose. Early reports from Israel on the effectiveness of the third dose have been published^11-16^, and demonstrate a marked decrease in new infections and specifically in severe cases. Following these reports, other countries recommended a third dose of the vaccine. While early data demonstrated that the receipt of a third BNT162b2 vaccine dose increased vaccine effectiveness to approximately 93-95%^11-14^, only limited real-world data are currently available for the BNT162b2 third vaccine dose safety or its effect on immunogenicity.

Here we present real-world data on vaccine effectiveness, safety and immunogenicity within a large-scale cohort of health care workers (HCW) in a large tertiary center in Israel, the Sheba HCW COVID Cohort.

## Results

The study includes three arms (Figure 1): (i) the immunogenicity arm, where serological tests of samples from vaccinated HCW at three time-points were assessed and compared: after receiving second dose, before receiving the third dose and after receiving the third dose; (ii) the vaccine effectiveness arm, where incidence of SARS-CoV-2 infections among HCW with two vaccine doses given at least 5 months previously was compared to incidence among HCW with three doses; (iii) the safety arm, where adverse events among those vaccinated with the third dose by September 2, 2021, were assessed via an electronic questionnaire.

**Figure 1:**
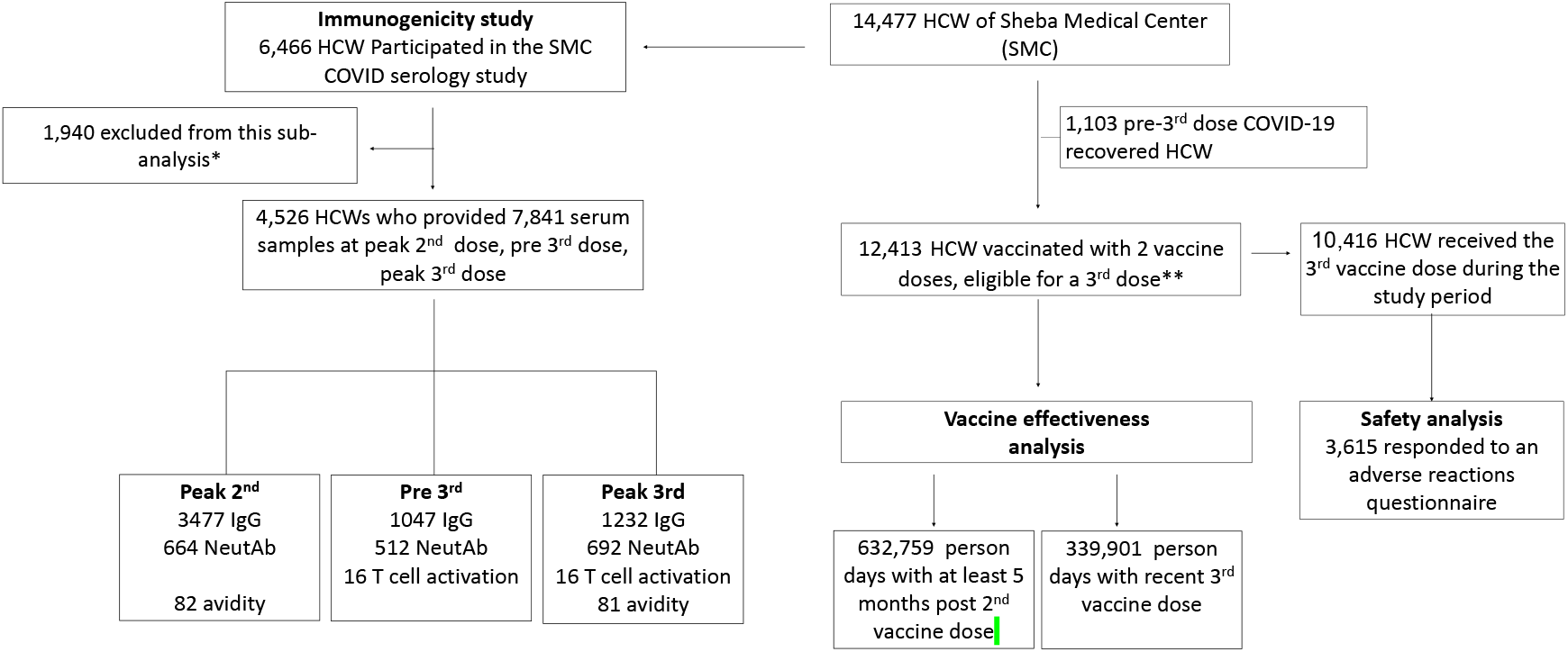
Study profile. The BNT162b2 vaccinated Sheba health care workers (HCW) cohort used for the vaccine effectiveness, safety and serology analyses following the second and third vaccination. HCW=health care work. IgG=immunoglobulin G. NeutAb=neutralizing antibodies.

### Immunogenicity data following the 3^rd^ BNT162b2 vaccine dose

Of 4,526 HCW eligible for the study, 1047 had serum samples from both pre- and post-third dose time-points (up to 45 days before, as well as 14-45 days after the third dose). IgG and neutralizing antibody levels at these two time points were tested for 1047 and 512 HCW, respectively. A 31-fold (95% confidence interval [CI], 28-34) and 41-fold (95% CI, 39-42) increase in IgG and neutralizing levels, respectively, was observed following the third vaccine dose (Figure 2a-b). Administration of the third dose also resulted in a minor, but significant increase (P=0.025) in T cell activation, which was tested in 16 HCW (Figure 2c).

**Figure 2:**
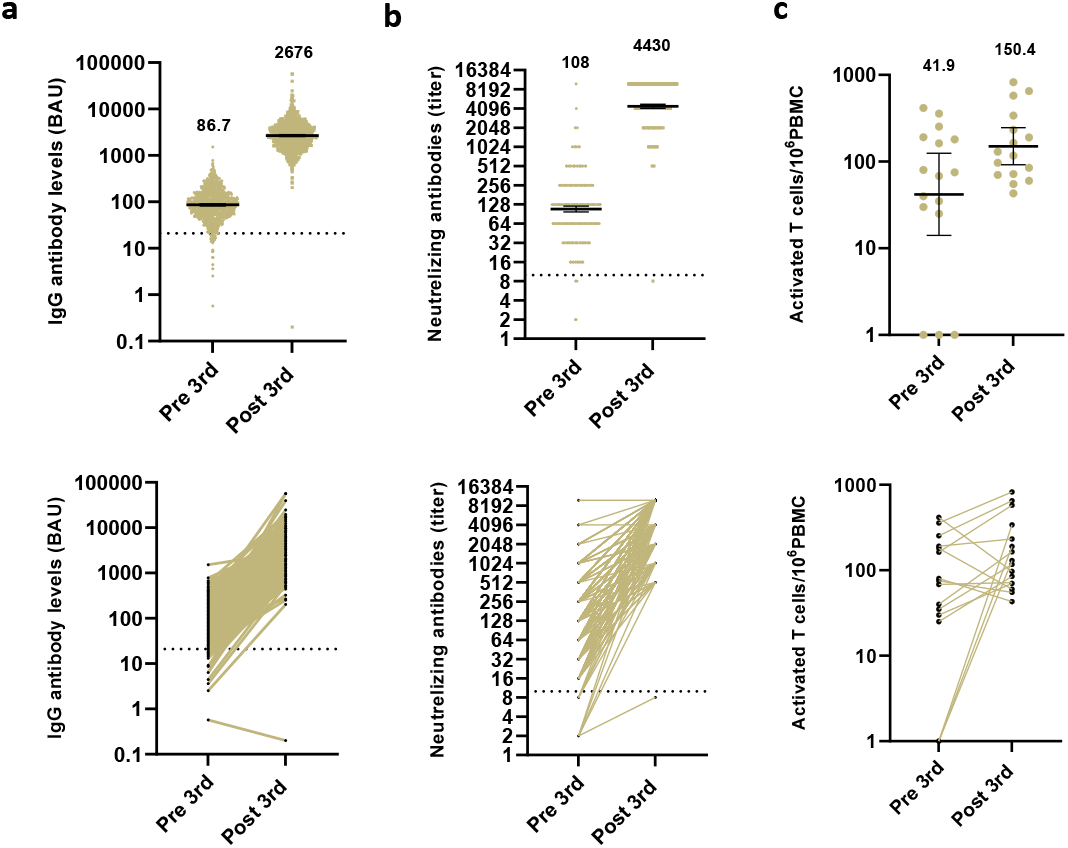
Immunogenicity pre and post 3rd vaccine dose. Scatter plot and before-after analysis of IgG antibodies **(a)**, neutralizing titers **(b)** and number of activated T cells **(c)** in HCW 45 days or less before or 14-45 days following the third vaccine dose. The dotted black line indicates the cutoff level of positive antibodies and neutralizing concentrations. Solid black lines indicate GMT with 95%CI. GMT of each time point is indicated. BAU=binding antibody units.

To investigate any added effect of the third vaccine dose on humoral response we compared IgG and neutralizing antibodies at their peak levels following the second vaccine dose (post-2^nd^) to that of the third dose (post-3^rd^). IgG and neutralizing antibody results were available for 3,477 and 664 HCW, respectively, after second vaccine dose and for 1,232 and 692 HCW after their third vaccine dose. A linear mixed model was used to examine the differences in immunogenicity across age, sex and number of co-morbidities, comparing post-second dose IgG levels and neutralizing antibody titers. The estimated geometric mean titer (GMT) for IgG following the second dose, in BAU was 1586 (95% CI, 1458-1709) and for IgG following the third dose, it was 2745 (95% CI, 2641-2853). Thus, a 1.7-fold (95% CI, 1.6-1.9) increase in IgG levels occurred following the third dose in comparison to following the second dose. Neutralizing antibody levels following the second and third vaccine doses were 646 (95% CI, 589-709) and 3948 (95% CI, 3735-4191), respectively (Figure 3a-b). The neutralizing titer following the third dose was thus 6.1-fold (95% CI, (5.5-6.8) greater than that of the second dose. Since both quantity and the strength of interaction of antibodies are important for neutralization, we tested IgG avidity post -2^nd^ and post 3^rd^ dose in 81 HCW randomly selected. While a 61.1% (95% CI, 56.1 to 66.7) avidity was observed following second dose, a substantially higher avidity of 96.3% (95% CI, 94.2 to 98.5) was found after the third dose (Figure 3c). No substantial differences in avidity at post-2nd and 3rd dose were observed between HCW 60 years old or older and below 60 years old (Figure S1).

**Figure 3:**
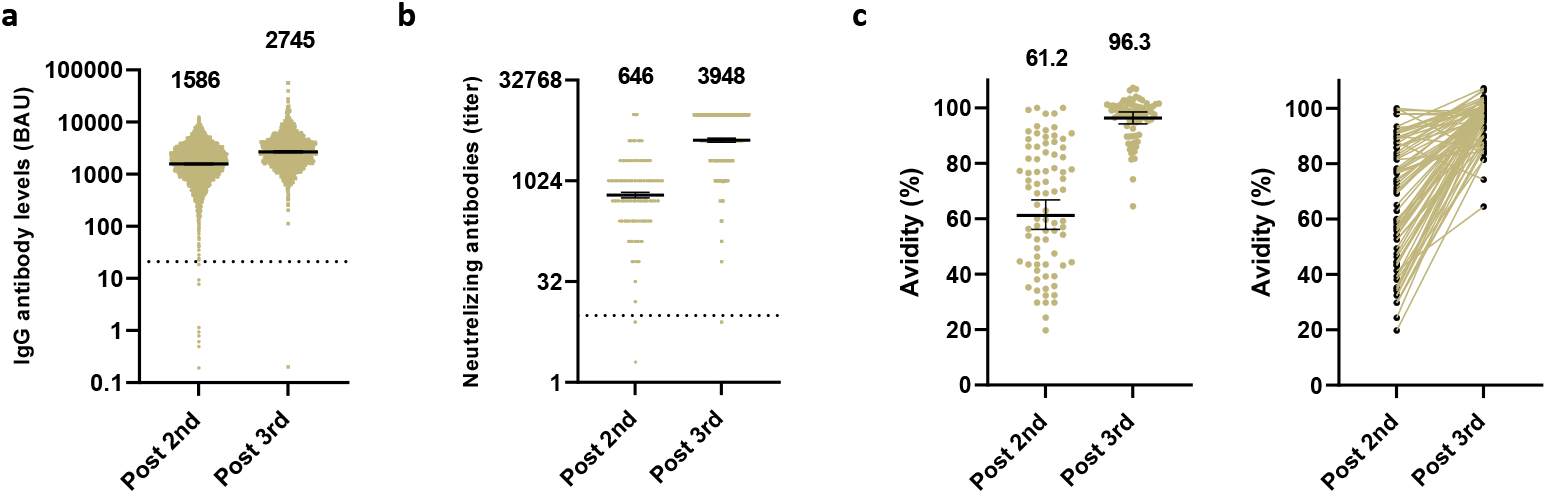
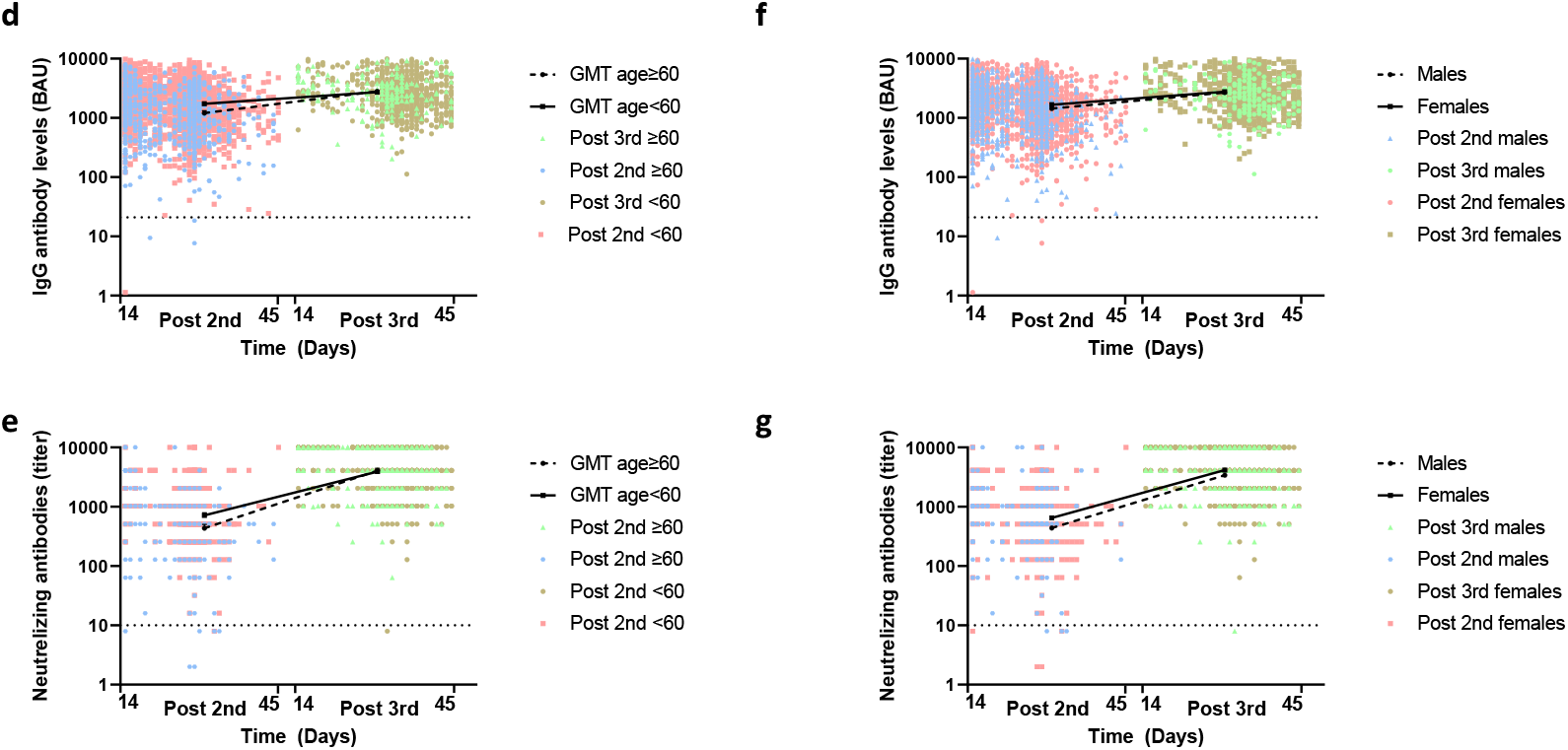
Humoral response 14-45 days after the second and third vaccine doses. **(a-c)** Scatter plot analysis of IgG antibodies **(a)** and neutralizing titers **(b)** and avidity scatter plot and before-after analysis **(c)** in HCW after the 2^nd^ (post 2^nd^) and third (post 3^rd^) vaccine doses. The dotted black line indicates the cutoff level of positive antibodies and neutralizing concentrations. Solid black lines indicate GMT with 95%CI. GMT of each time point is indicated. BAU=binding antibody units. **(d-g)** Expected GMT of IgG (d,f) and neutralizing antibodies (e,g) at post 2^nd^ and post 3^rd^ according to age group (d,e) and sex (f,g). Dots represent individual observed serum samples. The dashed line in each panel indicates the cutoff for diagnostic positivity. ⍰ bars indicate 95% confidence intervals.

Following the second dose, lower IgG and neutralizing titers were associated with older age, male sex and the presence of at least one (for IgG) or two (for neutralizing antibodies) coexisting conditions while higher IgG titers were associated with BMI of 30 or higher (obesity) as compared with a BMI of less than 30 (see Table 1 and Supplementary Materials, Section 4.3 for details). The third dose elicited a 1.41 (95% CI, 1.27 to 1.58) and 1.19 (95% CI, 1.06 to 1.34) fold increased expression of IgG antibodies and 1.66 (95% CI, 1.32 to 2.08) and 1.33 (95% CI, 0.91 to 1.95) fold increased expression of neutralizing titers, in HCW 60 years old and older and in HCW with two or more co-morbidities, respectively, compared with younger HCW and HCW with no morbidities. Concomitantly, following the third dose, differences in IgG levels between older and younger persons, gender and between those with and without comorbidities were much reduced and no longer statistically significant (see Table 1 and Supplementary Materials, Section 4.3 for details); higher levels of IgG antibodies continued to be associated with a BMI of 30 or greater following the third dose. Neutralizing antibody levels following the third dose were lower in males than females, but did not differ substantially according to age group, BMI group or number of comorbidities (Table 1).

**Table 1.**
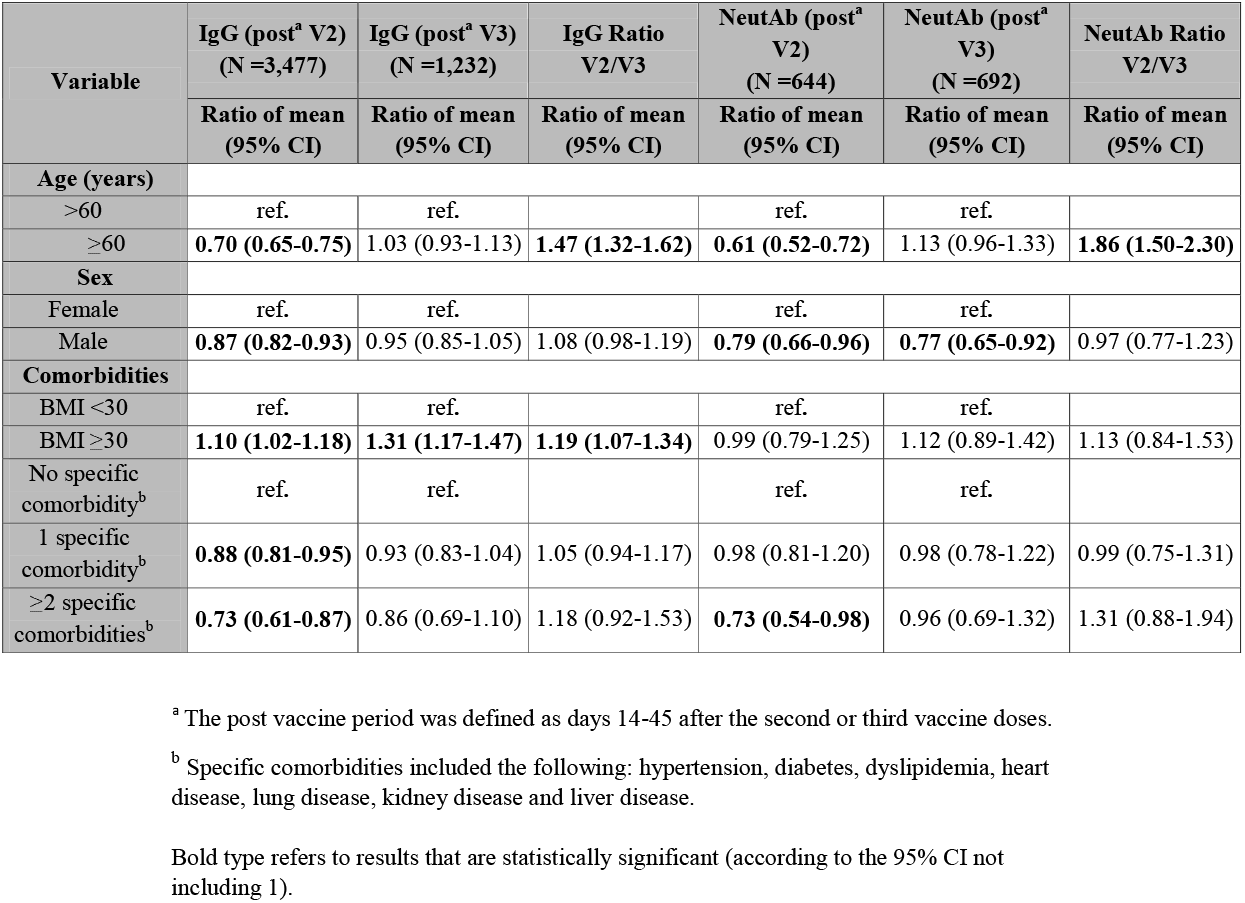
Mixed model analysis of variables associated with IgG and NeutAb titers following second and third vaccine doses.

| Variable | IgG (post<sup>a</sup> V2)<br>(N =3,477) | IgG (post<sup>a</sup> V3)<br>(N =1,232) | IgG Ratio<br>V2/V3 | NeutAb (post<sup>a</sup><br>V2)<br>(N =644) | NeutAb (post<sup>a</sup><br>V3)<br>(N =692) | NeutAb Ratio<br>V2/V3 |
| --- | --- | --- | --- | --- | --- | --- |
|  | Ratio of mean<br>(95% CI) | Ratio of mean<br>(95% CI) | Ratio of mean<br>(95% CI) | Ratio of mean<br>(95% CI) | Ratio of mean<br>(95% CI) | Ratio of mean<br>(95% CI) |
| **Age (years)** |  |  |  |  |  |  |
| >60 | ref. | ref. |  | ref. | ref. |  |
| ≥60 | **0.70 (0.65-0.75)** | 1.03 (0.93-1.13) | **1.47 (1.32-1.62)** | **0.61 (0.52-0.72)** | 1.13 (0.96-1.33) | **1.86 (1.50-2.30)** |
| **Sex** |  |  |  |  |  |  |
| Female | ref. | ref. |  | ref. | ref. |  |
| Male | **0.87 (0.82-0.93)** | 0.95 (0.85-1.05) | 1.08 (0.98-1.19) | **0.79 (0.66-0.96)** | **0.77 (0.65-0.92)** | 0.97 (0.77-1.23) |
| **Comorbidities** |  |  |  |  |  |  |
| BMI <30 | ref. | ref. |  | ref. | ref. |  |
| BMI ≥30 | **1.10 (1.02-1.18)** | **1.31 (1.17-1.47)** | **1.19 (1.07-1.34)** | 0.99 (0.79-1.25) | 1.12 (0.89-1.42) | 1.13 (0.84-1.53) |
| No specific<br>comorbidity<sup>b</sup> | ref. | ref. |  | ref. | ref. |  |
| 1 specific<br>comorbidity<sup>b</sup> | **0.88 (0.81-0.95)** | 0.93 (0.83-1.04) | 1.05 (0.94-1.17) | 0.98 (0.81-1.20) | 0.98 (0.78-1.22) | 0.99 (0.75-1.31) |
| ≥2 specific<br>comorbidities<sup>b</sup> | **0.73 (0.61-0.87)** | 0.86 (0.69-1.10) | 1.18 (0.92-1.53) | **0.73 (0.54-0.98)** | 0.96 (0.69-1.32) | 1.31 (0.88-1.94) |

<sup>a</sup> The post vaccine period was defined as days 14-45 after the second or third vaccine doses.
<sup>b</sup> Specific comorbidities included the following: hypertension, diabetes, dyslipidemia, heart disease, lung disease, kidney disease and liver disease.
Bold type refers to results that are statistically significant (according to the 95% CI not including 1).

### Correlation between IgG and neutralizing antibodies one month after the third dose

We next assessed whether the increase in avidity affected the correlation between IgG and neutralizing antibodies following the third vaccine dose. A similarly strong correlation was observed following the second dose (Spearman’s rank correlation of 0.62) as well as following the 3^rd^ dose (Spearman’s rank correlation of 0.61) (Figure S3).

### Vaccine effectiveness of the third dose

For this analysis 12,290 naïve HCW eligible for a third vaccine dose, for whom data was available, were included. These HCW contributed together 632,759 person days to the two-dose cohort and 339,901 person days to the three-dose cohort. In total, 407 HCW were found positive on PCR testing, 368 in the two-dose cohort and 39 in the three-dose cohort. The crude SARS-CoV-2 breakthrough incidence rate in the two-dose cohort was therefore 5.8 per 10,000 days at risk compared to 1.1 per 10,000 days in the three-dose cohort. After adjustment for gender, age, and time (weekly period), estimated vaccine effectiveness of the third dose relative to two doses was 85.6% (95% CI, 79.2-90.1%) (Table 2).

**Table 2:** Vaccine effectiveness of third dose vs. two outdated doses among 12,413 HCW.

|  | **Two-dose cohort\*** | **Three-dose cohort\*\*** | **Rate Ratio (RR)**<br>**(95% CI)** | **Relative VE (%)**<br>**100 (1 – 1/RR)**<br>**(95% CI)** |
| --- | --- | --- | --- | --- |
| **Cases** | 368 | 39 |  |  |
| **Person/day exposure** | 632,759 | 339,901 |  |  |
| **Crude Breakthrough Infection Rate/10,000PD** | 5.82 | 1.15 | 5.07<br>(3.64-5.07) | 80.3<br>(72.6-85.8) |
| **Adjusted Breakthrough Infection Rate/10,000PD** | 6.63 | 0.95 | 6.96<br>(4.82-10.05) | 85.6<br>(79.2-90.1) |
| * Second dose of vaccine given at least 5 months previously, no third dose |  |  |  |  |
| ** Third dose given at least 10 days previously |  |  |  |  |

### Adverse events (AE)

Of 8,337 HCW who were sent the electronic questionnaire, 3611 responded. The difference between those who responded to the questionnaire and those who did not are summarized at supplementary Table S1c. Figures S3a and S3b summarize the local and systemic reactions reported among responders.

The proportion of females among responders was significantly higher (p<0.0001) and the median age of the responders group was significantly lower (51.7, IQR 39.4-66.5, vs. 55.9, IQR 41.6-68.5, in non-responders). We thus stratified AE by gender and age group.

Local reactions were very common, with nearly all (95%) young females (age<60) and two thirds (68%) of the older male group reporting local reactions, mostly pain at the injection site. Systemic reactions, were also frequently reported by young females, with 76% reporting any systemic reaction, including fatigue and myalgia, and 19% reporting fever. Yet, only 31% of the older males reported any systemic reaction and only 3% reported fever (**Figure 4**, Sup Figs S3). Only two HCW required hospital admission due to symptoms that occurred in proximity to the receipt of the third vaccine dose; one suffered from a migraine with sensory loss and was hospitalized for 2 days and the other had unexplained hyponatremia and was discharged from the hospital after 4 days.

**Figure 4:**
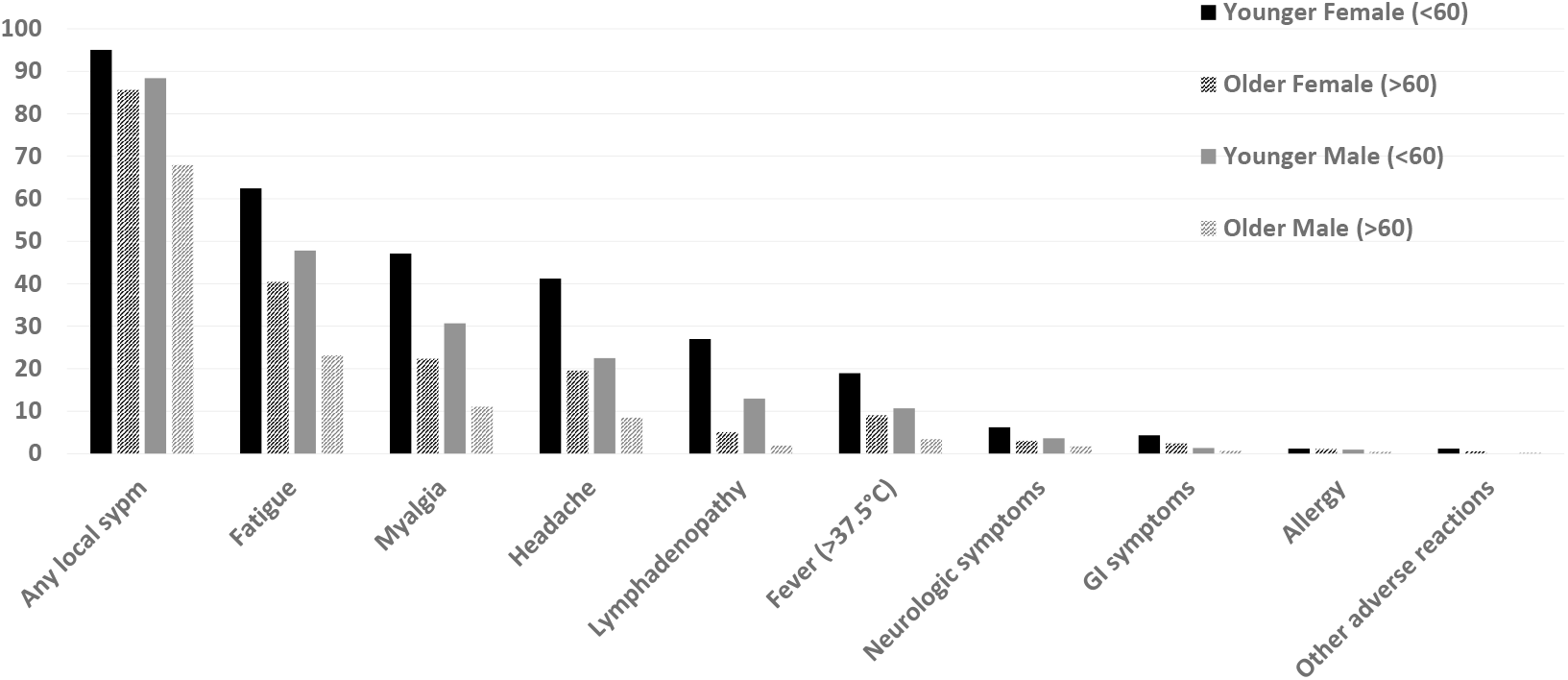
Local and systemic reactions reported following the third vaccine dose. Local symptoms include local pain (reported by 3,058 of 3,611 HCW), swelling (n=685), redness (n=445) and itching (n=11). Neurologic symptoms include paresthesia (n=138) and facial nerve palsy (n=17). Other adverse reactions include abnormal lab results such as elevated CRP, elevated TSH, hyponatremia, leukopenia (n=1 each); and other symptoms included herpes labialis (n=3) and report of new-onset psoriasis (n=1).

## Discussion

In this study we found that the humoral response generated by the third BNT162b2 vaccine dose is substantially superior over the response to the second dose, resulting in overall increased IgG levels, avidity and neutralizing antibody titers, eliminating the differential lower IgG and neutralizing levels observed in older and morbid populations. With mostly self-limiting adverse reactions, the third vaccine dose boosted vaccine effectiveness which was diminished 5-6 months following the second vaccine dose.

Accumulating data regarding immunogenicity demonstrate a significant increase in antibody and neutralizing levels in the first weeks following two BNT162b2 vaccine doses^17^ which decline over a period of at least six months with IgG antibodies decreasing at a consistent rate and neutralizing antibody titers plateauing after three months^9^. In conjunction with this finding, vaccine effectiveness was shown to be 90-95% in the first months and rapidly waning in all age groups with time following the second vaccine dose ^3,7,8^. Antibody and neutralizing titers were both demonstrated to be correlated with protection from infection^10,18,19^ suggesting that immunogenicity levels observed following the second dose can serve as a marker for immunity. With the administration of the third vaccine dose, key questions arising are whether antibody and neutralizing response after the third dose as well as immunogenicity kinetics will be similar to those of the second dose. Our results show that while a significant, yet small ∼1.7-fold increase in IgG antibody levels was observed, there was a ∼6-fold increase in neutralizing titers comparing with those after the second dose. This discrepancy between antibody levels and neutralization response is most likely due to the significant increase in the strength of interaction between IgG antibodies and SARS-CoV-2 antigen (avidity) observed following the third dose compared to that observed after the second dose. Interestingly, the increase in avidity following the third dose did not alter the correlation between IgG antibodies and neutralizing levels. Thus, our results demonstrate that the third vaccine dose elicits IgG affinity maturation which specifically impacts neutralization capacity. This suggests that lower IgG antibody levels will be required to maintain high neutralizing titers, and as a result, immunity may be sustained for a longer period of time following administration of the third vaccine dose, despite a similar rate of IgG waning. Accordingly, the high avidity antibodies generated by the third vaccine dose, may induce higher neutralizing protection against VOC such as the newly emerging Omicron.

Using a mixed model, we analyzed the association of age, sex, and coexisting conditions with immunogenicity one month after the second and third doses. Separately, we also investigated the immune response before and after the third dose. Consistent with our previous results^9^, a significantly lower antibody response was found among older HCW, males and HCW with two or more co-morbidities, one month after the second dose. Interestingly, HCW 60 years old and older and HCW with two or more co-morbidities had increased reaction following the third compared to younger and HCW with no morbidities, respectively. As a consequence, no significant difference was observed between old and young populations as well as between HCW with and without co-morbidities, one month after the third vaccination indicating that these vulnerable populations mount an immune response similar to that of healthy HCW. Indeed, a similar phenomenon of increased response to vaccination in older HCW was observed following the second BNT162b2 vaccine dose ^17^. It is interesting to speculate that while generation of a primary immune response may be hampered in vulnerable individuals, their secondary response is intact and able to compensate following sequential booster doses.

The relative VE of the third dose, of 85.6% measured in this study, comparing incidence rate among third dose recipients to that among those who were eligible for a third dose, but did not receive it, is slightly lower than three other observational studies from Israel, that reported VE of 88%-92% ^12,13,16^. The meticulous follow-up of Sheba HCW allowed us to examine the effectiveness of the third dose against infection even in asymptomatic and very mild symptomatic cases which are usually not identified in observational studies. As a result, we believe our data more accurately reflects the VE of symptomatic and asymptomatic individuals. Furthermore, our VE data was generated in the same cohort used for conducting the immunogenicity assays, thus enabling us to better associate VE with immune response. Continued monitoring of the Sheba HCW cohort will allow us to assess delta VOC correlates of protection and VE for Omicron and other future VOC.

Reactogenecity in our study was actively pursued and therefore we believe it did not miss adverse events, particularly not serious ones. Yet, our estimations of proportion of individuals reporting AE may be overestimated due to reporting bias. In our study, most third vaccine dose recipients reported mild to moderate and transient local and systemic reactions. The rate of adverse events here is similar to findings from several studies which monitored the safety of one and two doses of BNT162b2 administration among healthy individuals^20,21^. It is also similar to a recent study that examined the safety of a third dose of COVID-19 vaccine among vaccine recipients in the US using a self-monitoring surveillance system^22^. Overall, these results suggest that the reactogenicity to the third dose is frequent, yet limited to mild local and systemic events and not different from the adverse events identified in the first and second vaccine doses.

This study was conducted on HCW and therefore does not represent the general population. However, the continuous monitoring of this cohort even before the administration of the first vaccine dose on Dec 2020 allows us to thoroughly examine the impact of the BNT162b2 vaccine using repeated measurements on the same population which results in obtaining valuable immunogenicity data across ages, gender and co-morbidities. It is important to keep in mind that the chief VOC circulating during the study period was Delta and therefore VE can be different against other VOC that may emerge such as the newly discovered Omicron variant. Nevertheless, our immunogenicity data can be of great importance once vaccine penetration and correlates of protection levels are determined.

Taken together, our immunogenicity, VE and safety data, clearly demonstrate that the third BNT162b2 vaccine dose, given at least five months following the second dose, safely boosts protection from SARS-CoV-2 infection by significantly inducing broad humoral and cellular responses. The antibodies generated as a result of this booster dose are of high avidity and as such are superior and will most probably protect from infection also vulnerable populations significantly longer than second dose generated antibodies.

## Methods

### Ethical Statement

The protocol was approved by the Institutional Review Board of the Sheba Medical Center (SMC) and written informed consent was obtained from all study participants.

#### Study setting and period

The Sheba HCW COVID Cohort study, is an ongoing prospective cohort study following the SMC HCW which has been conducted since vaccination rollout first began on December of 2020^9-11,17^. SMC is the largest tertiary medical center in Israel, with 1,600 beds and 14,479 HCW, including employees, students, volunteers, and retired personnel. Between December 2020 and July 2021 a total of 95% of eligible HCW received two doses of the BNT162b2 vaccine. On July 28, 2021 the Israeli ministry of health decided to administer a third vaccine dose to individuals aged 60 years or older, on Aug 15 to younger HCW who received their second vaccine dose more than five months prior and on August 29, this decision was expanded to general population. Of all SMC HCW, 12,243 received two COVID-19 vaccine doses by May 2021, did not acquire SARS-CoV-2 by July 28, 2021 and were eligible to receive the third dose which was offered at SMC.

This study includes immunogenicity data from Feb, 2021 until Oct, 2021. The VE sub-study took place during the fourth surge of SARS-CoV-2 infections in Israel which was predominated by the delta variant of concern (VOC), between July 8 and October 1, 2021. Safety data was collected between Aug 7 and Sep 2, 2021.

#### Study design and population

The Sheba serology study, initiated before the rollout of the first COVID-19 vaccine dose and recruited 6,466 HCW, consisted of monthly serological follow-up. Here, we included HCW who fulfilled the following criteria: (i) 18 years or older, COVID-19-naïve, i.e., no history of SARS-CoV-2 infection determined by previous positive PCR, positive anti-S IgG before receiving the first dose or positive anti-N IgG at any time point, (ii) had available sera 14-45 days after second (Post 2^nd^) or/and (iii) had available sera 45 days or less before the third (Pre 3^rd^) or/and (iv) had available sera 14-45 days after third (Post 3^rd^) dose. For subjects who had more than one eligible sample in the Post 2^nd^ or Post 3^rd^ period, the sample with highest IgG levels were included in the analysis. For subjects who had more than one eligible sample in the Pre 3^rd^ dose period, the sample closest to receipt of the 3^rd^ vaccine dose was chosen. Baseline characteristics of the study population are presented in Table S1a-c.

Neutralizing antibody assays were performed on a selected subgroup that included higher proportions of persons with risk factors of interest, such as an age of 65 years or older and coexisting conditions. Criteria for the selection of participants for the neutralizing antibody subgroup are listed in Supplementary Methods 2.

For the vaccine effectiveness study, all HCW without history of SARS-CoV-2 infection by June 15, 2021, who received the second vaccine dose by May 1st and were eligible to receive the third dose between July 29 and October 2nd were included. Data on PCR testing methods are provided in Supplementary Methods Section.

Data on age and sex were available for all study participants. A computer-based questionnaire about demographic characteristics and coexisting conditions was sent electronically to all serology-study participants. The questionnaire and definitions of the study variables are provided in Tables S2 and S3.

#### Serology Assays

Samples from vaccinated participants were tested before receipt of the third dose using the SARS-CoV-2 Receptor Binding Domain (RBD) IgG assay (Beckman-Coulter, CA, U.S.A.), or after receipt of the third dose using the SARS-CoV-2 IgG II Quant (Abbott, IL, USA) test. These commercial tests were performed according to manufacturer’s instructions. To present all IgG Antibody levels in Binding Antibody Units (BAU) per the World Health Organization (WHO) standard measurements we imputed the Abott-based BAU values from the Beckman-Coulter assay results, based on an independent sample of individuals with both Abbott BAU and Beckman-Coulter levels (see Supplementary Methods 4.1).

SARS-CoV-2 Pseudo-virus (psSARS-2) Neutralization Assay was performed as described^17^ using a green fluorescent protein (GFP) reporter-based pseudotyped virus with a vesicular stomatitis virus (VSV) backbone coated with SARS-CoV-2 spike (S) protein generously provided by Dr. Gert Zimmer (Institute of Virology and Immunology (IVI), Mittelhäusern, Switzerland). The level of detection for IgG are and NeutAb are above 21.4 and 8, respectively. Additional information about antibody testing is provided in Supplementary information.

Avidity was determined by avidity ELISA using urea as a chaotropic reagent. information about avidity testing is provided in Supplementary information.

#### T-cell activation

Enzyme-linked immune absorbent spot (Elispot) was used as described^11^ to measure antigen-specific T cells that secrete Interferon-γ following vaccination. Additional information about T-cell activation is provided in Supplementary information.

#### Adverse event active surveillance

All HCW who received the 3rd vaccine dose before September 2nd were sent a short electronic questionnaire regarding side effects of the third vaccine dose. They were asked about various localized and systemic side effects, the duration of these symptoms and whether they required medical care or hospitalization. Additionally, HCW and their treating physicians were encouraged to report any serious adverse event or hospitalization. To identify reporting bias of those answering the questionnaire we compared demographic variables of responders and non-responders and accordingly stratified the outcome by under- or over-represented sub populations.

### Statistical analysis

#### Imputation of Abbott-based BAU values from Beckman-Coulter test values

Binding antibody levels measured in BAU were available only following receipt of the third dose. Before the receipt of the third dose binding antibodies were assessed using the Beckman-Coulter RBD assay. To make comparisons of before and after the third dose, Abbott-based BAU values were imputed from the Beckman-Coulter assay levels on the basis of an independent sample of individuals who had both measurements. For details of the imputation, see Supplementary Materials, Section 4.1. To account for the extra uncertainty due to using imputed values, bootstrap methods were used to calculate confidence intervals. See below.

#### Comparison of pre- with post-third vaccine antibody levels

All IgG antibody levels were analyzed on the natural logarithmic scale. Pre-third vaccine neutralizing antibody levels were compared to post-third vaccine levels, and the difference expressed as a ratio between geometric means, with the 95% confidence interval based on matched pre- and post-samples.

Pre-and post-third vaccine IgG levels were compared in the same way, but the confidence interval was based on taking bootstrap samples of both the imputation sample and the sample of HCW with pre- and post-third vaccine levels. See Supplementary Methods, Section 4.2 for more details.

#### Comparison of post-second with post-third vaccine antibody levels

This comparison included three subgroups of HCW: those with antibody levels measured after the second dose only, those with antibody levels measured after the third dose-only and those with measurements at both time points. Both neutralizing antibody and IgG levels at these time-points were compared using linear mixed models. Each individual’s level was modeled as a random effect, and time-point (post third vaccine versus post second vaccine) was modeled as a fixed effect. Individuals’ characteristics were included as fixed-effect adjusting covariates, and included gender, age group (<60y, ≥60y), body mass index (<30, ≥30, missing), and number of comorbidities (0, 1, ≥2, missing), where the comorbidities considered were hypertension, diabetes mellitus, dyslipidemia, heart disease, lung disease, kidney disease and liver disease. Interaction terms between each adjusting covariate and the time-point variable were also included. Parameter estimates from the model were then used to compare post-third dose to post-second dose levels, overall and in subgroups of adjusting covariates, as well as comparing post-second dose levels according to covariates and post-third dose levels according to covariates.

Results are expressed as ratios of geometric means. For neutralizing antibodies, p-values and 95% confidence limits were calculated directly from the model output. For binding antibodies, they are based on bootstrapping of both the imputation sample and the HCW sample. See Supplementary Materials, Section 4.3 for more details.

#### Vaccine effectiveness

We investigated vaccine effectiveness of the third vaccine dose relative to two doses given at least 5 months previously, for the period July 1^st^ to October 2^nd^. Two cohorts of HCW were defined: the “two-dose” cohort including those eligible for the third dose (previously uninfected and having received their second dose at least 5 months previously) but not having received it, and the “three-dose” cohort including those who had received their third dose (10 or more days previously). Individuals in the two-dose cohort exited that cohort on the day that they were diagnosed with a positive PCR test or on the day they received the third dose. Individuals entered the three-dose cohort on the tenth day following receipt of the third dose and exited on the day that they were diagnosed with a positive PCR test. Follow-up terminated on October 2^nd^, 2021.

Incidence rates were analyzed using a Poisson Regression model. The follow-up period was divided into weekly periods, for each period the number of positive diagnoses and the number of follow-up days was calculated for each cohort, subdivided into four subgroups according to gender and age (<60y and ≥60y). From this model, we estimated the ratio of incidence rates in the two cohorts (third dose versus two doses only), adjusted for gender, age and period. Confidence intervals (95%) were calculated based on model standard errors of the estimated log ratio.

#### Safety data

To determine the representability of those who responded to the AE questionnaire, we compared demographic characteristics of responders to non-responders using Chi Square and 2-sample student t-test.

#### Graphical presentation

Scatter plots of IgG and neutralizing antibody levels since the receipt of the second and third doses were created with the use of GraphPad Prism software, version 9.0 (GraphPad Software).

Correlations between IgG and neutralizing antibody levels for each period were assessed by Spearman’s rank correlation. Paired pre- and post-third vaccine dose avidity, neutralization and T cell activation were compared using the Wilcoxon signed-rank test. Statistical analysis was performed using SAS software, version 9.4 (SAS Institute), and the linear mixed-model analyses were performed using R software, version 3.6.2 (R Foundation for Statistical Computing).

## Data Availability

All data produced in the present study are available upon reasonable request to the authors

## Acknowledgements

This study was funded by internal funds of the Sheba Medical Center. YL is a recipient of the Nehemia Rubin Excellence in Biomedical Research, The TELEM Program of Chaim, Sheba Medical Center. We greatly acknowledge Tal Levin, Ravit Koren, Shiri Kats-Likvornik, Osnat Halpern, Yara Kanaaneh, Sabha Abosiam, Alex Aydenzon, Maayan Chiara Atias-Golbus for their devoted work in the laboratory, the IPC team for the extensive epidemiologic investigation performed, Yael Beker-Ilani, Etti Rozner, and Efrat Steinberger for coordinating the study.

