## Supplementary Appendix for "Superior immunogenicity and effectiveness of the 3rd BNT162b2 vaccine dose"

This appendix has been provided by the authors to give readers additional information about their work.

**Supplementary Methods 1 - PCR testing**

Hospital personnel were tested in several scenarios: upon every symptom suspected to be COVID-19, following exposure to a positive COVID-19 contact (hospital or community contacts), as part of a “return to work” protocol during the end of isolation period following exposure or disease.

For quantitative RealTime-PCR (qRT-PCR), nasopharyngeal swabs were placed in 3mL of universal transport medium (UTM) or viral transport medium (VTM). Test was performed according to manufacturers' instructions on various platforms: Allplex™ 2019-nCoV (Seegene, S. Korea), NeuMoDx™ SARS-CoV-2 assay (NeuMoDx™ Molecular, Ann Arbor, Michigan), Xpert®, Xpress SARS-CoV-2 (Cepheid, Sunnyvale, CA, USA).

**Supplementary Method 2- Inclusion criteria for selecting the neutralizing antibody subgroup**

1. Age ≥65
2. Body mass index ≥30
3. Pregnancy
4. Allergy
5. Hypertension
6. Diabetes
7. Dyslipidemia
8. Heart disease
9. Lung disease
10. Kidney disease
11. Liver disease
12. Autoimmune disease
13. Immunosuppression

Additionally, 50% of healthy health care workers were randomly selected for the neutralizing antibody subgroup.

**Supplementary Methods 3- Immunogenicity**

**SARS-CoV-2 Pseudovirus (psSARS-2) Neutralization Assay**

SARS-CoV-2 Pseudo-virus (psSARS-2) Neutralization Assay was performed using a propagation-competent VSV-spike similar to the one previously published^2^ which was kindly provided by Gert Zimmer, University of Bern, Switzerland and shown to be highly correlative to authentic SARS-CoV-2 virus micro-neutralization assay. Following titration, 100 focus forming units (ffu) of psSARS-2 were incubated with 2-fold serial dilution of heat inactivated (56°C for 30 min) tested sera. After incubation for 60 min at 37°C, virus/serum mixture was transferred to Vero E6 cells that have been grown to confluency in 96-well plates and incubated for 90 min at 37°C. After the addition of 1% methyl cellulose in dulbecco's modified eagle's medium (DMEM) with 2% of fetal bovine serum (FBS), plates were incubated for 24hr and 50% plaque reduction titer was calculated by counting green fluorescent foci using a fluorescence microscope (EVOS M5000, Invitrogen). Sera not capable of reducing viral replication by 50% at 1 to 16 dilution or below were considered non- neutralizing. For clear presentation non- neutralizing samples were marked as a titer of 2.

**Avidity**

All samples were subjected to an "in house" RBD-IgG ELISA as previously described^1^ with the addition of 6M urea or PBS for 10 min for each sample. Briefly, a 96 well microtiter Polysorb plate (Nunc, Thermo, Denmark) was coated overnight at 4°C with 50μl per well of 1μg/ml of RBD antigen. After blocking with 5% skimmed milk at 25°C for 60 minutes, serum samples diluted 1:100, 1:400 and 1:1000 with 3% skimmed milk, were added to antigen coated wells. The plate was incubated at 25°C for 120 minutes, and following washing each sample was incubated either with the addition of 6M urea or PBS for 10 min.After washing, a goat anti-human IgG horseradish peroxidase (HRP) conjugate (Jackson ImmunoResearch, PA, USA Code: 109-035-088) (diluted 1:15000) was added to each well for 60 min. After washing, incubation of TMB Substrate Solution (Abcam) for 5 min and the addition of stop solution (2N HCl), the OD of each well was measured at 450nm using a micro-plate reader (Sunrise, Tecan). Avidity index was calculated as the ratio (in percentage) between sample OD with 6M urea and sample OD with PBS.

**Peripheral blood mononuclear cells (PBMC) isolation**

To assess for T-cell response, PBMC were isolated by density gradient centrifugation using UNI-SEP+ (Novamed). Plasma was collected and spun at 1000 × g for 20 min to remove platelets before collection of PBMC. Following one wash with PBS and one wash with 4Cell Nutri-T-Medium (Sartorius), cells were re-suspended in 4Cell Nutri-T-Medium and counted using the Countess II Cell counter (Invitrogen).

**IFN-γ ELISpot assay**

IFN- γ -secreting cells were enumerated using Elispot IFN-γ kits (IFN-γ kit, AID Autoimmun Diagnostika GmbH, Strassberg, Germany) according to manufacturer instructions. For antigen stimulation, 50 μl of SARS-CoV-2 peptide pools (S-complete, Miltenyi Biotech) were used. Test medium was used as negative control and Phytohaemagglutinin (PHA) was used as positive control. IFN-γ-secreting cells frequency was quantified using the AID ELISpot Reader (Strassberg, Germany). The unspecific background (mean SFU from negative control wells) was subtracted from experimental readings.

**Supplementary Methods 4 – Statistical Methods**

**4.1 Imputation of Binding Antibody Units based on Beckman-Coulter Assay Results**

We developed a method for imputing Abbott IgG levels from Beckman-Coulter IgG levels using data on 215 selected serum samples, taken from individuals who had not received a booster dose and were not included in the HCW cohort, and were measured by both methods. We fitted a cubic polynomial regression model where log (to the base e) of IgG measured in BAU units by the Abbott kit was regressed on the log (to the base e) of IgG measured by the Beckman-Coulter kit, its squared value, and its cubed value. The fitted regression equation, using the glm procedure in R, was:

logIGG_Abbott =

4.506 + 0.6634 × logIGG_Beckman - 0.0852× (logIGG_Beckman)^2^ + 0.0403× (logIGG_Beckman)^3^

**Output from the glm procedure in R**

```
Coefficients:
              Estimate Std. Error t value Pr(>|t|)
(Intercept)   4.506257   0.039815 113.180  < 2e-16 ***
logBeck       0.663432   0.042540  15.595  < 2e-16 ***
I(logBeck^2) -0.085175   0.020513  -4.152 4.78e-05 ***
I(logBeck^3)  0.040341   0.007083   5.695 4.10e-08 ***
---
Signif. codes:  0 ‘***’ 0.001 ‘**’ 0.01 ‘*’ 0.05 ‘.’ 0.1 ‘ ’ 1

Residual standard error: 0.4377 on 211 degrees of freedom
Multiple R-squared:  0.9187,	Adjusted R-squared:  0.9176
```

A simple linear model has an R-squared of 0.905, compared to 0.919 for the cubic polynomial. One can see from the figure below that the cubic polynomial fits the data much better at the lower and upper ends of the scale.

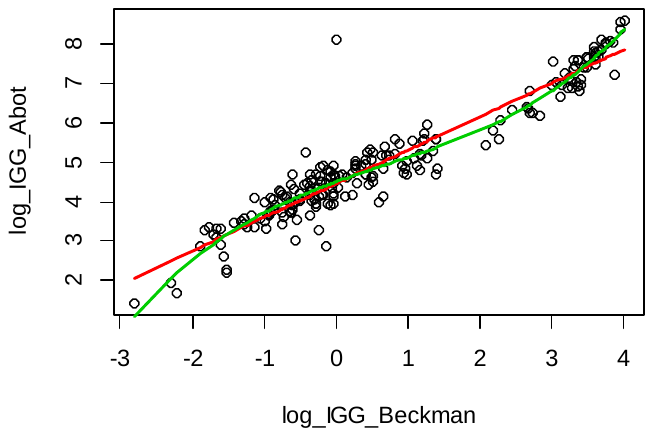

The regression equation shown above was used to impute the values of Abbott BAU for samples taken before the third (booster) dose.

**4.2 Comparison of pre- with post-third vaccine IgG levels**

We included only participants who were measured before (Beckman-Coulter) and after (Abbott) the 3^rd^ vaccine and excluded the very few participants who had an Abbott kit measurement at the pre-3rd vaccine. The number of participants included in the analysis was 1047. We used bootstrapping of both the imputation sample and the pre-post 3^rd^ vaccine sample (2000 repeats) to obtain confidence intervals for the mean change on the log scale from pre- to post-vaccination. Having established that the bootstrap distribution of estimates appeared very symmetric, we used the percentile method to estimate the confidence interval. In addition to the average change, we computed the geometric mean IgG levels at pre and post vac3 for the total sample and stratified by either gender or age group. The confidence intervals of these means were also obtained by the bootstrap percentile method.

The results are given in the table below. The “Ratio” row is the ratio of the post-vaccine IgG level to the pre-vaccine level. The GM column gives the geometric means. In the tables stratified by gender and age group, the Ratio column compares geometric means of females versus males, or individual less than 60y versus those aged 60y and over. Levels for females, both pre and post were higher than for males. For those aged under 60y pre-vaccine levels were higher than for those 60y and over, but post-vaccine levels for the two subgroups were similar.

**Total sample (IgG scale)**

```
           GM   2.5%   97.5%
preV3     86.7   79.9   95.1
postV3  2676.6 2550.8 2797.4
Ratio     30.9   28.2   33.6
```

**By Gender**

```
	Female    2.5%   97.5%    Male    2.5%   97.5% Ratio 2.5% 97.5%
          GM                       GM
preV3    89.95   82.61   98.52   75.41   65.84   86.52 1.19  1.06  1.36
postV3 2758.73 2631.72 2889.29 2382.97 2082.23 2683.53 1.16  1.02  1.33
Ratio    30.67   27.88   33.46   31.60   27.82   36.02 0.97  0.86  1.10
```

**By age group**

```
         <60y    2.5%   97.5%     60+    2.5%    97.5% Ratio 2.5% 97.5%
          GM                      GM
preV3    88.91   81.76   97.64   77.39   67.54   88.50  1.15 1.01  1.30
postV3 2667.75 2530.89 2801.79 2717.90 2458.42 2995.35  0.98 0.88  1.10
Ratio    30.00   27.27   32.77   35.12   30.64   40.26  0.85 0.75  0.97
```

**4.3 Comparison of post-second with post-third vaccine IgG levels**

We included subjects who were measured either after the 3^rd^ (Abbott) or the 2^nd^ (Beckman-Coulter) vaccination. There were 3787 subjects included: 922 were measured after both second and third vaccinations, 2555 were measured after the 2^nd^ vaccination only, and 310 were measured after the third vaccination only. We estimated the difference between the post-third vaccine value and the post-second vaccine value, on the log Abbott BAU scale, using a linear mixed model, where each person’s level was modeled as a random effect, and post-3^rd^ dose versus post-2^nd^ dose was modeled as a fixed effect. Individuals’ characteristics were included as fixed-effect adjusting covariates, and included gender, age group (<60y, ≥60y), BMI (<30, ≥30, missing), and number of comorbidities (0, 1, ≥2, missing), where the comorbidities considered were hypertension, diabetes, dyslipidemia, heart disease, lung disease, kidney disease and liver disease. Interaction terms between each adjusting covariate and the time-point variable were also included. BMI and comorbidity data were missing in 23% of the sample, and, in order to include these persons in the analysis, we included a missing category for each of these variables. We used bootstrapping of both the imputation sample and the post-vaccinations sample (2000 repeats) to obtain confidence intervals, stratifying the latter sample into three strata: those measured after second dose only, those measured after third dose only, and those measured at both time-points. Confidence limits were calculated using the bootstrap percentile method, having found that the bootstrap distributions were very symmetric.

The estimates of the parameters in the model, obtained from the statistical package in R software, version 3.6.2 (R Foundation for Statistical Computing) are shown below.

```
                 est     2.5%    97.5%
(Intercept)  1828.967 1671.614 1984.065
v3              1.479    1.349    1.628
male            0.874    0.821    0.928
agec60+         0.702    0.649    0.754
bmi30           1.099    1.020    1.178
bmi30miss       0.960    0.739    1.229
ndis1           0.883    0.808    0.953
ndis2+          0.729    0.607    0.870
ndismiss        0.993    0.770    1.302
v3:male         1.082    0.979    1.193
v3:agec60+      1.468    1.323    1.622
v3:bmi30        1.193    1.071    1.343
v3:bmi30miss    1.268    0.813    2.002
v3:ndis1        1.054    0.942    1.173
v3:ndis2+       1.182    0.917    1.529
v3:ndismiss     0.837    0.517    1.315
```

Aside from the intercept term which is on the BAU scale, each coefficient is a multiplicative factor associated with that covariate. Reference categories for the covariates are female, aged <60y, BMI <30, and no comorbidities. The parameters of interest are v3 and its interaction with the covariates. For example, v3 measures the factor increase in post vaccine 3 level compared to post vaccine 2 level for a woman aged <60y, with BMI <30 and no comorbidities (48% increase). The interaction term v3:agec60+ measures how different is the change in the IGG levels from post-vaccine 2 to post-vaccine 3 between those aged 60+ and those aged <60y (47% increase). If the value 1 is within the 95% CI the effect of the covariate is regarded as non-significant at the 5% level. One can see that the multiplicative change between post vaccine 2 and post vaccine 3 levels is larger for those aged over 60y, and for those with BMI greater than or equal to 30.

The terms male, agec60+, bmi30, ndis1 and ndis2 give the ratios of the IgG levels at post vaccine 2 between the subgroup and its reference subgroup. For example, males had IgG levels post vaccine 2 that are 89.5% of the levels of females, and the confidence limits exclude the value 1, meaning that the female levels were statistically significantly higher than those of males. Post vaccine 2 IgG levels appeared to be higher also in older persons and those who were obese.

The following table gives the corresponding ratios for the post vaccine 3 measurement.

```
            est  2.5% 97.5%
male      0.946 0.849 1.046
agec60+   1.030 0.933 1.131
bmi30     1.312 1.169 1.470
bmi30miss 1.217 0.818 1.869
ndis1     0.930 0.829 1.042
ndis2+    0.861 0.686 1.095
ndismiss  0.831 0.538 1.247
```

It can be seen that the levels of IgG post-vaccine 3 differ statistically significantly according to BMI (with obese patients having higher levels), but not according to gender, age, or comorbidity.

We also examined the geometric mean IgG levels at post-vaccine2 and post-vaccine 3 of the full sample, and by gender and age group, adjusted for all covariates. They were obtained by estimating the expected IGG (using the estimated coefficients of the fixed effects from the model) of all participants in post-vaccine periods and averaging their expected levels. The confidence intervals are obtained by the bootstrap procedure.

The results are given in the Table below:

**Total sample (Abbott IgG scale)**

```
             est    2.5%   97.5%
postV2   1585.58 1457.51 1708.77
postV3   2744.60 2640.62 2852.53
Dif         1.73    1.60    1.90
```

**By Gender**

```
       Female   2.5%  97.5%   Male   2.5%  97.5%
postV2 1659.7 1519.3 1796.1 1399.2 1282.6 1523.1
postV3 2788.2 2680.3 2904.4 2628.7 2398.0 2880.1
```

**By age group**

```
          <60   2.5%  97.5%    60+   2.5%  97.5%
postV2 1733.1 1588.0 1873.3 1166.5 1054.3 1278.9
postV3 2732.8 2618.7 2854.9 2785.8 2571.3 3028.6
```

Similar methods were used for analyzing neutralizing antibody levels post-vaccine 2 and post-vaccine 3, but confidence intervals were calculated from model-based standard error estimates, since no imputation was necessary for these levels.

**4.4 Vaccine effectiveness – detailed results**

As explained in the Methods section of the main text, we used a Poisson regression model to evaluate the effectiveness of the third dose (10 or more days after administration) compared to two doses given at least 5 months previously. The parameter estimates from the model, obtained from the *glm* procedure in R software, are shown below.

The log incidence rate ratio for the third dose versus two doses is given by the coefficient for the “booster” variable (see the row in bold type). The rate ratio estimate is therefore 0.144 (exponent of

-1.9396), which translates into a relative vaccine effectiveness of 85.6%.

The other rows show the relationship of the adjusting covariates, age, gender and period to the incidence rate. It can be seen that in this study, those aged 60 and above were at lower risk than younger workers. It can also be seen that the incidence surge reached its peak between 19^th^ August to 16^th^ September.

```
                  Estimate Std. Error z value Pr(>|z|)
Intercept)        -8.9715     0.3806 -23.573  < 2e-16 ***
booster           -1.9396     0.1877 -10.332  < 2e-16 ***
male               0.1103     0.1064   1.036  0.29999
agec60+           -1.3995     0.2251  -6.217 5.06e-10 ***
period2021-07-08   0.3363     0.4928   0.682  0.49500
period2021-07-15   0.3077     0.4928   0.624  0.53239
period2021-07-22   0.9072     0.4421   2.052  0.04018 *
period2021-07-29   1.0428     0.4340   2.403  0.01627 *
period2021-08-05   2.1045     0.3990   5.274 1.33e-07 ***
period2021-08-12   1.9514     0.4055   4.812 1.50e-06 ***
period2021-08-19   2.4689     0.4017   6.146 7.94e-10 ***
period2021-08-26   2.8778     0.3982   7.226 4.97e-13 ***
period2021-09-02   2.1148     0.4263   4.960 7.04e-07 ***
period2021-09-09   2.5192     0.4147   6.074 1.24e-09 ***
period2021-09-16   2.0482     0.4381   4.675 2.94e-06 ***
period2021-09-23   1.8266     0.4584   3.985 6.76e-05 ***
period2021-09-30   1.6362     0.5909   2.769  0.00562 **
```

Adjusted incidence rates were calculated by applying the above model parameters to calculate the expected incidence rates for each covariate profile, first under receipt of the third dose and then under no receipt of the third dose. A weighted average of these expected incidence rates was then taken, with weights equal to the person days at risk on each profile.

**Supplementary Table S1a – Baseline characteristics of the serology study population**

| **Variable** | **SMC serology study cohort included in the analysis (N=4,526)** | **NeutAb subgroup***  **(N=1,199)** |
| --- | --- | --- |
| **Gender, n (%)** |  |  |
| Female | 3298 (72.9) | 898 (74.9) |
| Male | 1228 (27.1) | 301 (25.1) |
| **Age, median (IQR)** | 46.95 (36.65-57.85) | 52.47 (40.37-65.39) |
| **Age, mean (+/-std)** | 47.74 (13.6) | 52.51 (14.5) |
| Age, n (%) |  |  |
| <60 | 3,579 (79.1) | 765 (63.8) |
| ≥60 | 947 (20.9) | 434 (36.2) |
| **BMI, mean (+/-std)** |  |  |
| **BMI, n (%)** | 3129 | 967 |
| <25 | 1,604 (51.3) | 493 (51) |
| 25-29.99 | 1,019 (32.6) | 312 (32.3) |
| 30+ | 506 (16.2) | 162 (16.8) |
| **Comorbidity, n (%)** | 3163 | 977 |
| Hypertension | 363 (11.5) | 154 (15.8) |
| Diabetes | 173 (5.5) | 65 (6.7) |
| Dyslipidemia | 208 (6.6) | 92 (9.4) |
| Heart disease | 91 (2.9) | 38 (3.9) |
| Lung disease | 107 (3.4) | 39 (4) |
| Kidney disease | 17 (0.5) | 6 (0.6) |
| Coagulation disorder | 61 (1.9) | 28 (2.9) |
| Liver disease | 23 (0.7) | 8 (0.8) |
| Autoimmune disease | 210 (6.6) | 107 (11) |
| **Specific comorbidities**, n (%)** |  |  |
| 0 | 2471 (78.1) | 685 (70.1) |
| 1 | 534 (16.9) | 218 (22.3) |
| ≥2 | 158 (5) | 74 (7.6) |

*NeutAb subgroup was defined as HCW with as at least one NeutAb assays during study follow-up. **Specific comorbidities included the following: hypertension, diabetes, dyslipidemia, heart disease, lung disease, kidney disease and liver disease. BMI=Body mass index; NeutAb=neutralizing antibodies

**Supplementary Table S1b – baseline characteristics of the vaccine effectiveness study population**

| **Variable** | **HCW eligible for a 3^rd^ vaccine dose from July 28^th^ to October 2^nd^ (N=12,243)** | **HCW who received a 3^rd^ vaccine dose by October 2^nd^ (N=10,332)** |
| --- | --- | --- |
| **Gender, n (%)** |  |  |
| Female | 8,542 (69.8) | 7,184 (69.5) |
| Male | 3,701 (30.2) | 3,148 (30.5) |
| **Age, median (IQR)** | 46.99 (35.12-62.82) | 49.37 (36.71-64.96) |
| Age, n (%) |  |  |
| ≤59.99 | 8,698 (71) | 6966(67.4) |
| ≥60 | 3,545 (29) | 3,366 (32.6) |

**Supplementary Table S1c – Adverse events questionnaire responders and non-responders**

| **Variable** | **Responders** | **Non responders** | **P value** |
| --- | --- | --- | --- |
| N | 3,611 (43.33%) | 4,718 (56.66%) |  |
| **Gender, n (%)** |  |  | < 0.0001 |
| Male | 996 (27.6%) | 1663 (35.2%) |  |
| Female | 2615 (72.4%) | 3055 (64.7) |  |
| **Age, median (IQR)** | 55.9 (41.6-68.5) | 51.7 (39.4-66.5) |  |
| Age, n (%) |  |  | < 0.0001 |
| <60 | 2,056 (56.9) | 3,039 (64.4) |  |
| ≥60 | 1,555 (43.0) | 1,679 (35.6) |  |

**Supplementary Table S2 – Serology study computer-based questionnaire**

|  | **Question** | **Answer1** | **Answer2** |
| --- | --- | --- | --- |
| **1** | What is your date of birth? |  | |
| **2** | What is your gender? | Male | Female |
| **3** | What is your current height in m? |  | |
| **4** | What is your current weight in kg? |  | |
| **5** | Did you perform an IgG assay before receiving the first dose of the vaccine? | Yes | No |
| **6** | Do you suffer from systemic hypertension (systolic blood pressure above 140) treated with medication? | Yes | No |
| **7** | Do you have dyslipidemia (total cholesterol above 200 or LDL cholesterol above 160) treated with medication? | Yes | No |
| **9** | Do you suffer from an autoimmune disease treated with medication? | Yes | No |
| **10** | Do you have diabetes (HbA1C>6.5 or fasting blood sugar>126) treated with medication? | Yes | No |
| **11** | Do you suffer from heart disease for which you are pharmaceutically treated? | Yes | No |
| **12** | Do you suffer from lung diseases such as asthma, COPD, pulmonary fibrosis treated with medication/s? | Yes | No |
| **13** | Do you have any coagulation disorder resulting in hemorrhage or thrombosis treated with medication? | Yes | No |
| **14** | Are you immunosuppressed (organ transplant recepient, currently undergoing biologic therapy/ chemotherapy, treated with corticosteroids, underwent a splenectomy, or diagnosed with HIV)? | Yes | No |
| **15** | Have you ever had a serious allergic reaction (anaphylaxis) that required immediate treatment? | Yes | No |
| **16** | Do you have liver disease as cirrhosis, hepatitis, liver cancer, metabolic disorder? | Yes | No |
| **17** | Do you have kidney disease (creatinine>1.2 or GFR<60) treated with medication? | Yes | No |
| **18** | Are you pregnant (confirmed by a beta HCG blood test and ultrasound fetal heartbeats detection)? | Yes | No |

The questionnaire was reviewed and approved by the Institutional review board of the Sheba Medical Center**.**

IgG=Immunoglobulin G; BMI=Body mass index; Kg=kilogram. M=meter; LDL=low-density lipoproteins; HbA1C=hemoglobin A1C; COPD= chronic obstructive pulmonary disease; HIV=human immunodeficiency; GFR=Glomerular filtration rate.

**Supplementary Table S3- Variable Definitions**

| **Variable** | **Values** | **Definitions** | **Timing** |
| --- | --- | --- | --- |
| **Outcomes** | | | |
| Post 2^nd^ IgG | Continuous (BAU) | **Expected** SARS-CoV-2 Receptor Binding Domain (RBD) Immunoglobulin G (IgG) assay (Beckman-Coulter, CA, U.S.A.) | During the peak period (days 14-45 after the second vaccination) |
| Post 2^nd^ NeutAb | Continuous (50% titer) | SARS-CoV-2 Pseudo-virus (psSARS-2) Neutralization Assay | During the peak period (days 14-45 after the second vaccination) |
| Post 3^rd^ IgG | Continuous (BAU) | SARS-CoV-2 Receptor Binding Domain (RBD) Immunoglobulin G (IgG) assay (Abbott, U.S.A.) | During the peak period (days 14-45 after the third vaccination) |
| Post 3^rd^ NeutAb | Continuous (50% titer) | SARS-CoV-2 Pseudo-virus (psSARS-2) Neutralization Assay | During the peak period (days 14-45 after the third vaccination) |
| **Variables** | | | |
| Sex | Female/male | As defined in SMC' files | Current |
| Age | Continuous (years) | As defined in SMC' files | At second vaccine dose |
| BMI | Categorical: <30, ≥30 | BMI was calculated by weight (kg)/(height (m))^2^ according to the HCW answer to the questionnaire. | At second vaccine dose |
| Blood pressure disease | 0/1 | According to the HCW answer to the questionnaire: defined as systolic blood pressure above 140 treated with medication | At second vaccine dose |
| Dyslipidemia | 0/1 | According to the HCW answer to the questionnaire: defined as total cholesterol above 200 or LDL cholesterol above 160 treated with medication | At second vaccine dose |
| Autoimmune disease | 0/1 | According to the HCW answer to the questionnaire: defined as known autoimmune disease treated with medication | At second vaccine dose |
| Diabetes | 0/1 | According to the HCW answer to the questionnaire: defined as HbA1C>6.5 or fasting blood sugar>126 treated with medication | At second vaccine dose |
| Heart disease | 0/1 | According to the HCW answer to the questionnaire: defined as known heart disease treated with medication | At second vaccine dose |
| Lung disease | 0/1 | According to the HCW answer to the questionnaire: defined as defined as known lung disease treated with medication | At second vaccine dose |
| Coagulation disorder | 0/1 | According to the HCW answer to the questionnaire: defined as known hemorrhage or thrombosis disease treated with medication | At second vaccine dose |
| Allergy | 0/1 | According to the HCW answer to the questionnaire: defined as a serious allergic reaction (anaphylaxis) that required immediate treatment | During the life |
| Liver disease | 0/1 | According to the HCW answer to the questionnaire: defined as cirrhosis, hepatitis, liver cancer, metabolic disorder | At second vaccine dose |
| Kidney disease | 0/1 | According to the HCW answer to the questionnaire: defined as creatinine>1.2 or GFR<60) treated with medication | At second vaccine dose |
| Pregnancy | 0/1 | According to the HCW answer to the questionnaire: defined as confirmed pregnancy by a beta HCG blood test and ultrasound fetal heartbeats detection | At second vaccine dose |
| Specific comorbidities | Categorical: 0, 1, ≥2 | Count of comorbidities that were with significant lower antibodies titers compared to healthy people during the first 5 weeks after the first vaccine dose^3^:   - Hypertension - Diabetes - Dyslipidemia - Heart disease - Lung disease - Kidney disease - Liver disease | At second vaccine dose |

Abbreviations: NeutAb= neutralizing antibodies; EoS=end of study; IgG=Immunoglobulin G; S/CO=sample cutoff ratio; SARS-CoV-2=severe acute respiratory syndrome; BMI=Body mass index; Kg=kilogram. M=meter; HCW=health care worker; LDL=low-density lipoproteins; HIV=human immunodeficiency; GFR=Glomerular filtration rate; HbA1C=hemoglobin A1C.

**Supplementary Figure S1: Avidity 14-45 days after the second and third vaccine.** Scatter plot analysis of avidity levels in HCW equal or above 60 (>59, 29 individuals) or below 60 (<60, 52 individuals) years old. Solid black lines indicate GMT with 95%CI. Significance between younger and older populations were tested by Mann-Whitney test.

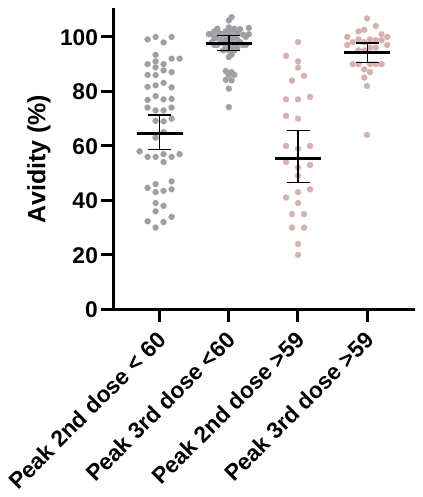

**Supplementary Figure S2: Correlation of IgG and Neutralizing antibodies.** The correlation was analyzed 14-45 days after the second and third vaccine dose.

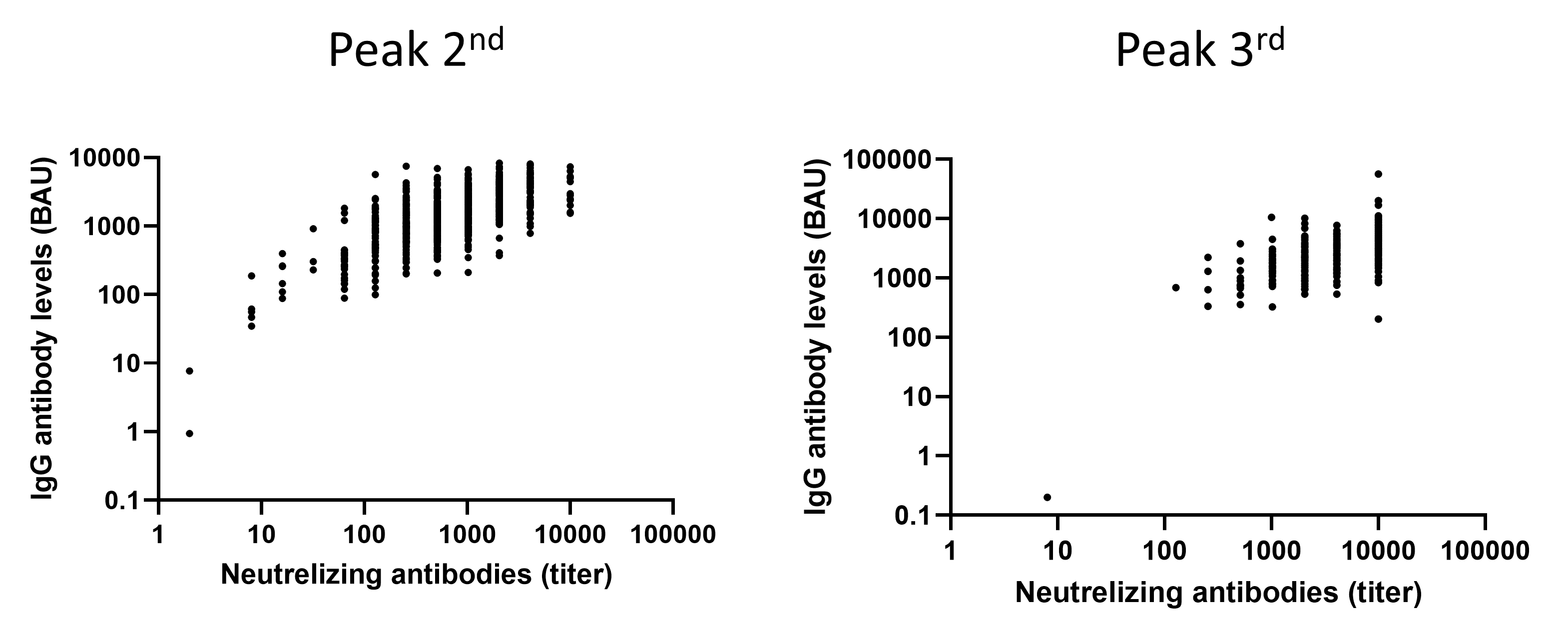

**Supplementary Figure S3: Local and systemic reactions reported following the third vaccine dose.** Local (a) and systemic (b) reactions with time and among ages and gender.

a. Local Symptoms

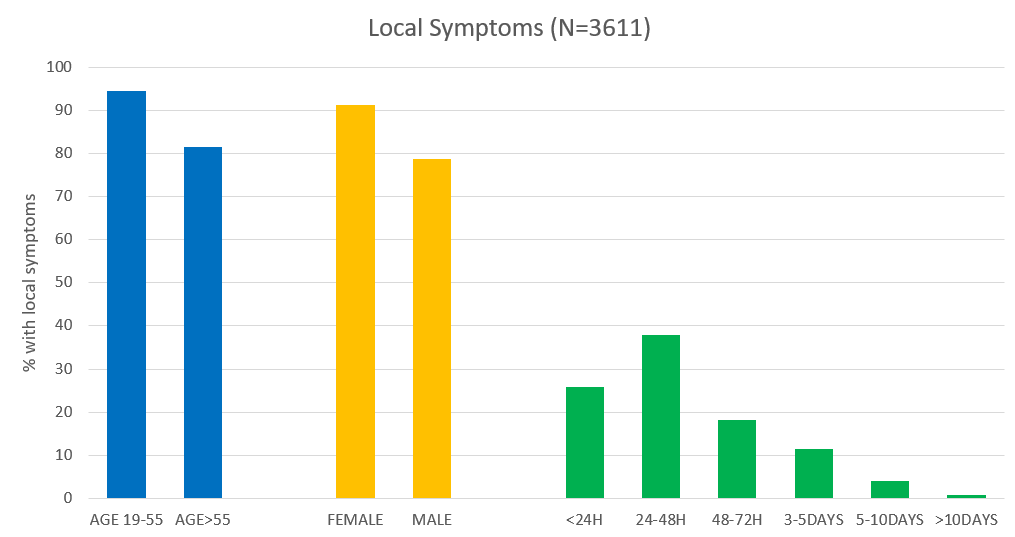

b. Systemic Symptoms

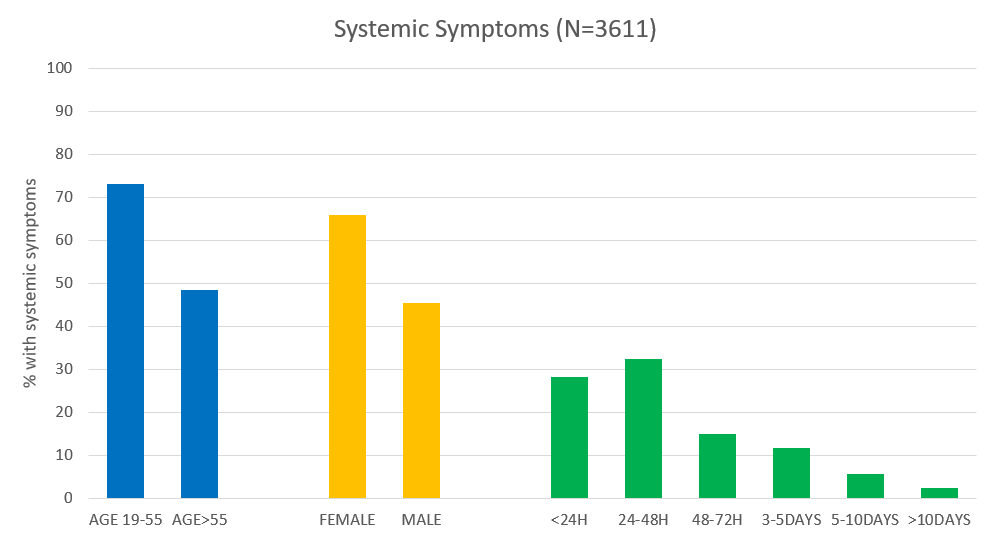
